# Mixed Methods Evaluation of the ‘*Caring for Providers to Improve Patient Experience*’ intervention

**DOI:** 10.1101/2023.10.12.23296648

**Authors:** Patience A. Afulani, Monica Getahun, Jaffer Okiring, Beryl A. Ogolla, Edwina N. Oboke, Joyceline Kinyua, Iscar Oluoch, Osamuedeme Odiase, Dan Ochiel, Wendy Berry Mendes, Linnet Ongeri

## Abstract

**Objective:** To assess the impact of the “Caring for Providers to Improve Patient Experience (CPIPE)” intervention, which sought to improve person-centered maternal care (PCMC) by addressing two key drivers—provider stress and bias.

**Methods:** CPIPE was successfully piloted over 6-months in two health facilities in Migori County, Kenya in 2022. The evaluation employed a mixed-methods pretest-posttest-non-equivalent-control-group design. Data are from surveys with 80 providers (40 intervention; 40 control) at baseline and endline, and in-depth-interviews with 20 intervention providers. We conducted bivariate, multivariate, and difference-in-difference analysis of quantitative data and thematic analysis of qualitative data.

**Results:** In the intervention group, average knowledge scores increased from 7.8(SD=2.4) at baseline to 9.5(SD=1.8) at endline for stress(p=0.001), and from 8.9(SD=1.9) to 10.7(SD=1.7) for bias(p=0.001); perceived stress scores decreased from 20.9(SD=3.9) to 18.6(SD=5.3)(p=0.019), and burnout from 3.6(SD=1.0) to 3.0(SD=1.0)(p=0.001); with no significant change in the control group. Qualitative data indicated CPIPE had impact at multiple levels. At the individual level, it improved provider knowledge, skills, self-efficacy, attitudes, behaviors, and experiences. At the interpersonal level, it improved provider-provider and patient-provider relationships leading to a supportive work environment and improved PCMC. At the institutional level, it created a system of accountability for providing PCMC and nondiscriminatory care; and collective action and advocacy to address sources of stress.

**Conclusion:** CPIPE impacted multiple outcomes in the theory of change leading to improvements in both provider and patient experience, including for the most vulnerable patients. These findings will contribute to global efforts to prevent burnout and promote PCMC and equity.

**Clinical trial registration:** ClinicalTrials.gov Identifier: NCT05019131 https://www.clinicaltrials.gov/study/NCT05019131?term=Afulani&checkSpell=false&rank=1

## Introduction

Person-centered maternal care (PCMC), which refers to responsive, respectful, and compassionate care, is an important dimension of quality of care and indicator of a human rights approach to care [1–4]. Yet, research continue to document poor PCMC across settings—manifesting as poor communication and lack of autonomy, and neglectful, disrespectful, and abusive treatment of women during facility-based childbirth [5–7]. This research also highlights inequities in PCMC with the most vulnerable groups having the worst experiences [6–10]. Two key drivers of poor PCMC and inequities in PCMC are provider burnout and bias [11–17].

Burnout, which is a state of physical, mental, and emotional exhaustion, is an important predictor of both poor patient and provider experience [18,19]. A key symptom of burnout, depersonalization, manifests as negativism, cynicism, and poor attitudes towards patients, leading to poor patient experiences [18,19]. Further, burnout is characterized by decreased professional efficacy, resulting in poor job satisfaction [20,21]. Burnout is caused by prolonged stress from exposure to stressors—environmental factors over which people have no control [22]. For maternal healthcare workers in low resource settings, these stressors are numerous: high workload, lack of basic resources, unsupportive work environments, limited competency to manage obstetric and newborn emergencies, and repeated trauma from avoidable morbidity mortality among others [23–25]. Provider burnout while previously high, has risen to crises levels since the COVID-19 pandemic [26,27], with particularly high rates among providers in Sub-Saharan Africa (SSA) [28–30]. In Kenya, over 80% of maternal health providers had moderate to high perceived stress and low to high burnout levels [31,32]. Further studies among maternal health providers in Kenya highlight provider stress and burnout as a key driver of poor PCMC [11,13,14,16].

Bias—perceiving and treating people differently according to their social group—on the other hand, leads to differential patient experiences [33,34]. Bias can be implicit (unconscious or unintentional) or explicit (conscious or intentional), and manifests as differing care based on patients’ attributes such as socio-economic status (SES), racial identity, ethnicity, and age. For example, lower SES patients often have more negative healthcare experiences than higher SES patients due to providers perceiving them to be less intelligent, compliant, and uninterested in promoting their health [35–37]. Providers also spend less time with low SES patients and provide insufficient information, contributing to distrust, poor adherence to treatment recommendations, and health disparities over time [38–40]. Studies in SSA, including Kenya have highlighted the role of provider bias in PCMC, demonstrating that healthcare workers provide differential care based on women’s appearances, perceptions of women’s ability to understand and be cooperative, and women’s ability to advocate for themselves and hold providers accountable [13–15,17].

Patient experience is thus intrinsically linked to provider experience. Provider stress and burnout contribute to negative patient experiences [11–13,16], while provider bias contributes to inequities in PCMC [13–15]. Further, research shows that people are more likely to be biased when stressed [41]. Yet, there are no documented evidence-based interventions to improve PCMC that address these underlying drivers in an integrated manner. To address this gap, we developed the *“Caring for Providers to Improve Patient Experience (CPIPE)”* intervention, focused on addressing provider stress and bias. CPIPE was designed iteratively guided by literature, behavior change theory—Social Cognitive Theory [42], Trauma Informed System framework [43], and the Ecological Perspective [44]—formative research, and continuous feedback from key stakeholders in Kenya. CPIPE has five strategies—provider training, peer support, mentorship, embedded champions, and leadership engagement. The training, which includes content on PCMC, stress management, dealing with difficult situations, and implicit and explicit bias, aims to increase provider knowledge, skills, and self-efficacy to impact attitudes and behaviors towards preventing burnout, mitigating bias, and improving PCMC. The remaining intervention strategies are intended to create an enabling environment for ongoing behavior change [45]. We piloted *CPIPE* to assess its feasibility, acceptability, and preliminary effectiveness. The intervention development and the implementation process are described elsewhere [46,47]. In this manuscript, we aim to assess the effect of the pilot intervention on key study outcomes. We hypothesized that CPIPE would enable providers to better manage stress to prevent burnout and increase providers’ awareness of their biases and motivation to avoid biased behaviors, which would improve both providers’ and patients’ experiences and mitigate the effects of their biases, thus reduce inequities in PCMC.

## Materials and Methods

### Study design and setting

We evaluated CPIPE using a pretest-posttest non-equivalent-control group design embedded within a mixed methods approach [48,49]. The study was conducted in Migori County in western Kenya from August 2021-July 2022. Migori County has a population of about 1.1 million, with a crude birth rate of 30.8 per 1000 [50,51]. It has eight sub-counties, each with a sub-county hospital and several health centers; provider/patient ratio is estimated at 32 nurses and four doctors per 100,000 people, respectively [52]. Most births (89.2%) occur in health facilities in the County [53]. The study setting is previously described [3,54].

In consultation with our community advisory board (CAB), we selected two high volume delivery facilities (facilities with more than 500 births per year)—Migori County Referral Hospital (MCRH) and Kuria West Sub County Hospital (Kehancha)—as intervention sites based on the number of providers available to participate. We then selected four other high volume delivery facilities as control facilities (Rongo, Ntimaru, Awendo, and Isibania sub-county hospitals). Allocation was not random, and more control sites were included because of the relatively fewer providers there compared to MCRH. The characteristics of study facilities are provided in table 1. All maternity providers—inclusive of clinical staff (nurses, midwives, doctors, clinical officers) and support staff (nurse aides, cleaners, etc.) from antenatal, intrapartum, and postnatal care units—in the study facilities were eligible to participate if they did not plan to relocate during the study period.

**Table 1:** Characteristics of Study facilities.

| Facility | Maternity bed capacity | Deliveries per year | Number of providers in maternity unit |  |  |  |
| --- | --- | --- | --- | --- | --- | --- |
|  |  |  | Doctors | Nurses/ midwives | Clinical officers | Support staff |
| MCRH | 100 | 4200 | 3 | 23 | 2 | 4 |
| Kehancha | 24 | 1300 | 1 | 7 | 0 | 3 |
| Rongo | 13 | 1440 | 1 | 9 | 2 | 3 |
| Ntimaru | 20 | 1200 | 0 | 9 | 2 | 1 |
| Awendo | 12 | 600 | 1 | 8 | 2 | 4 |
| Isbania | 12 | 600 | 0 | 7 | 2 | 3 |

### Intervention description

The CPIPE intervention strategies are summarized in Figure 1. The training content was first delivered through short didactic and interactive sessions over two days, with some content integrated into an emergency obstetric and neonatal care (EmONC) simulation to enable providers to apply concepts in the context of an emergency. This was followed by monthly refreshers facilitated by the embedded champions in their facility. We also created a *WhatsApp* group where resources were shared weekly with providers. The peer support groups were cadre-specific in-person groups based on the preferences of the providers, facilitated monthly by a peer leader. Groups discussed topics of their choice including debriefing on events at the maternity unit, discussing problems and brainstorming solutions; and engaging in stress management activities such as mindfulness exercises, singing, and dancing. The mentorship strategy was an onsite in-person mentorship model with intentional mentor-mentee pairing based on a survey, which identified areas for mentorship and individual needs and preferences. Embedded champions in each facility organized intervention activities, delivered training content during monthly refreshers, modeled behavior change, and provided support and feedback. A multi-level leadership engagement started during the planning phase in the form of a CAB with representation at the County, facility, and unit levels, as well as provider and patient representatives. The CAB, which met quarterly, provided input on the intervention strategies as well as the implementation and served as a place for high level advocacy for issues at the facility level. The intervention was delivered from November 2021-May 2022. The control group did not receive any intervention during the intervention period.

**Figure 1:**
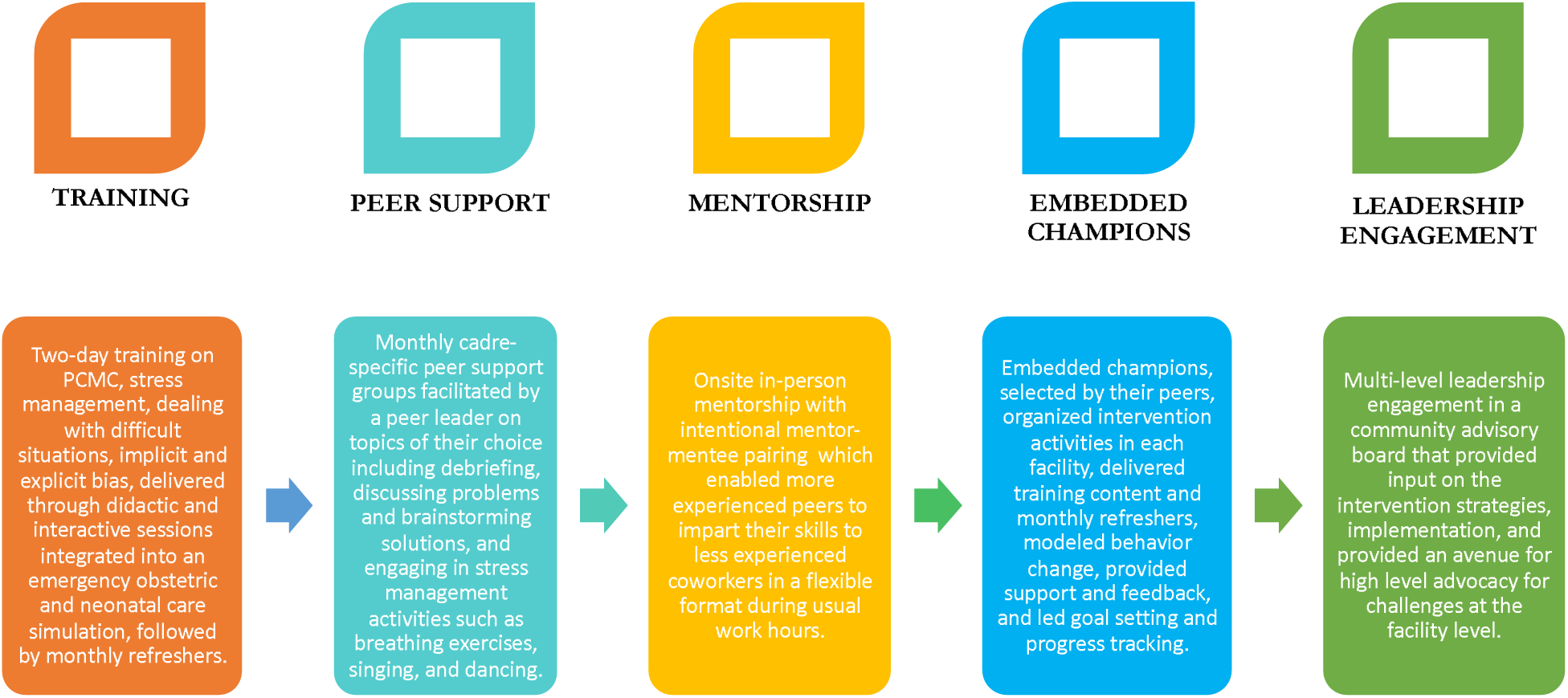
CPIPE Intervention Strategies.

### Data collection

We collected quantitative data through close-ended surveys with providers in the intervention and control groups before the launch of the intervention (baseline) and after the end of the intervention (endline); qualitative data were collected through in-depth interviews (IDIs) at endline. The intervention timing and data collection are shown in figure 2.

**Figure 2:**
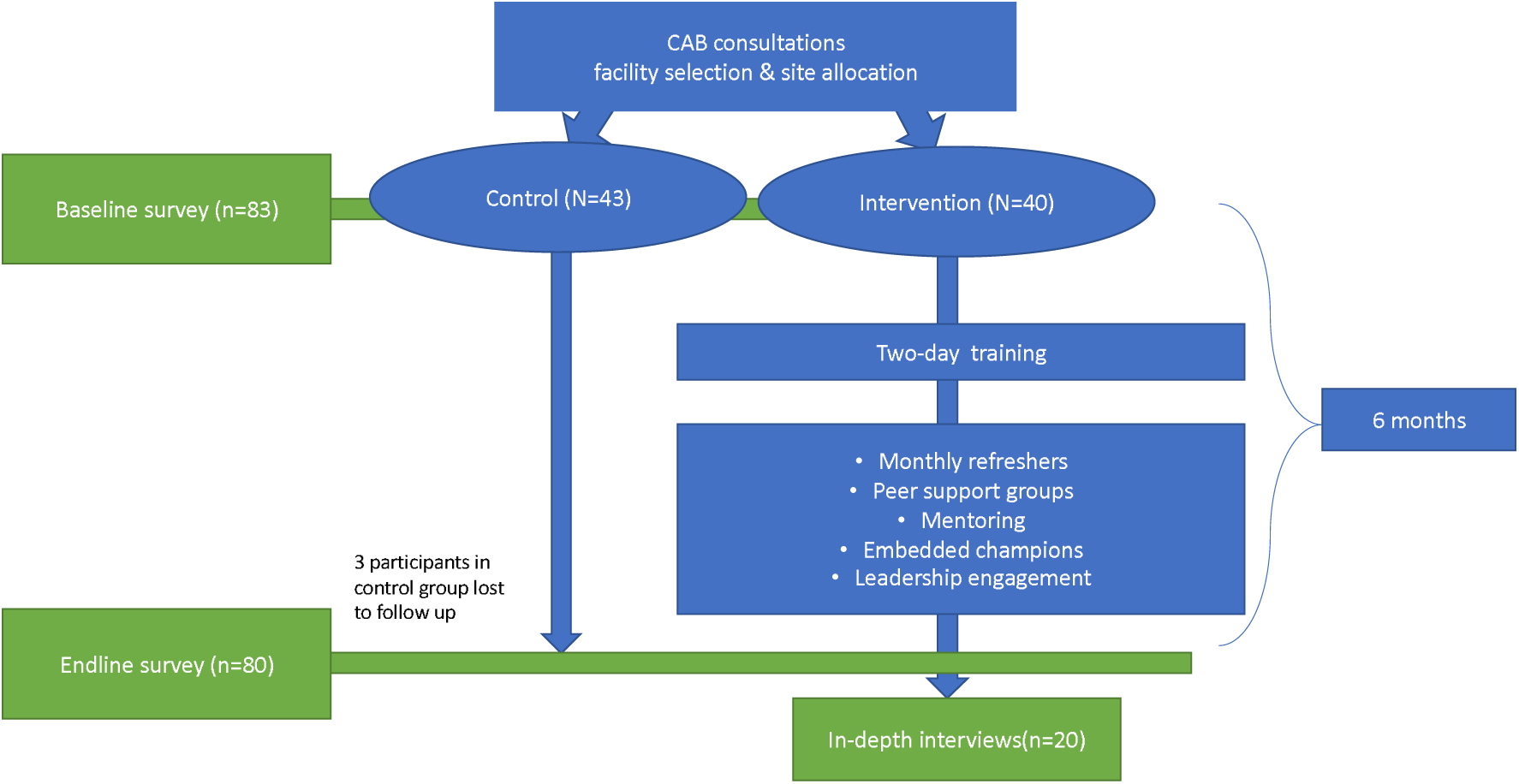
Flowchart of study activities.

### Quantitative survey

To recruit for the surveys, we first obtained a list of all providers in the maternity unit at each facility. Two female Kenyan research staff (BO and EN), trained in qualitative and quantitative research data collection, approached providers, explained the study, and obtained informed consent. All participants who were approached agreed to participate. Baseline surveys were conducted with 83 providers from August to October 2021, and follow-up surveys with the 80 participants from May to June 2022. Three participants from the baseline cohort were unavailable at endline (two moved outside the County and one on maternity leave). Sample size based on feasibility given pilot study. One-on-one structured surveys were conducted in English, at a private location in the facility, using tablets programmed with REDCap [55]. Interviews lasted 40 to 60 minutes.

### Qualitative

IDIs were conducted with a sub-set of 20 providers from the intervention sites who participated in the surveys, from July-September 2022, purposively sampled based on facility and position. BO and EN conducted the IDIs in the participant’s preferred language (English, Swahili, or Luo), using a semi-structured interview guide. Interviews were audio recorded and transcribed.

### Study ethics

Ethical approval was obtained from the Institutional Review Boards of the Kenya Medical Research Institute (SERU 3682) and the University of California, San Francisco (IRB number 17-22783), with additional approvals from the Kenya National Commission for Science, Technology and Innovation and the Migori County Commissioner and Director of Health. All participants provided written informed consent and received an incentive of 300 Kenyan shillings (∼$3) for each interview.

### Quantitative Measures

The primary outcomes were (1) stress knowledge, (2) implicit bias knowledge, (3) perceived stress, and (4) burnout. Secondary outcomes were (1) explicit bias, (2) implicit bias, (3) PCMC provision, and (4) physiologic measures of stress: heart rate variability (HRV) and hair cortisol levels. We used preexisting validated or new tools developed by our team to measure all outcomes described in Box 1. Additionally, we collected socio-demographic and job-related data to control for other factors which may explain the intervention outcomes including age, gender, marital status, parity, education, perceived social status, religion, position, years of work experience, workdays and hours, and prior training on stress management, bias, and patient-provider interactions.

#### Box 1

**Study outcome measures**

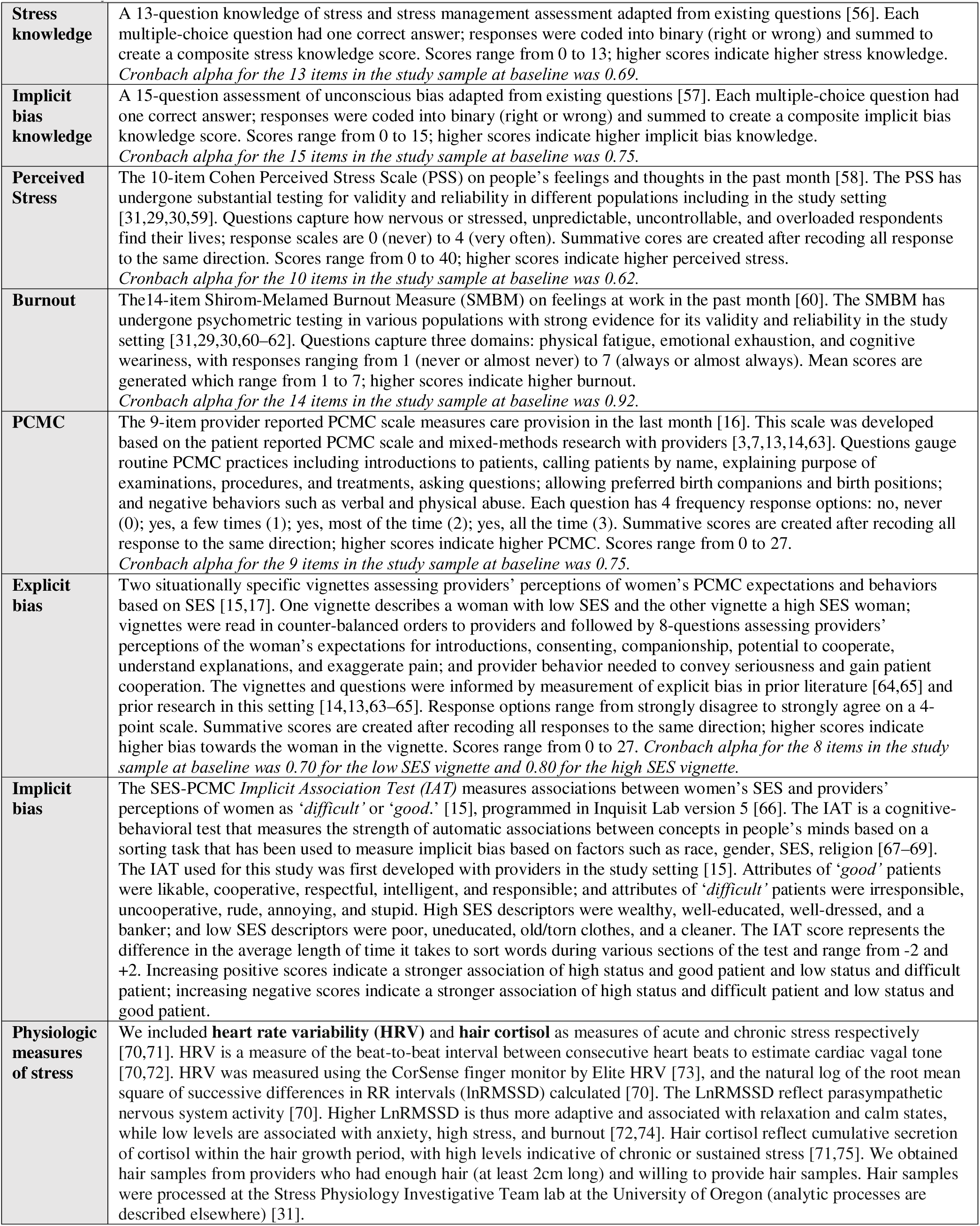

### Statistical analysis

We used descriptive statistics (means and proportions) to examine characteristics of the sample and distributions of the outcome variables. We then conducted factor analysis of the perceived stress, burnout, explicit bias, and PCMC items to assess construct validity and Cronbach’s alpha to assess internal consistency reliability, before generating summative scores used in subsequent analysis. For respondents missing responses, we used item-mean substitution to impute scale scores [76]. This was done for only two respondents with incomplete data on the explicit bias items. We tested whether there was a change in outcome measure scores from baseline to endline using Wilcoxon paired signed-rank test for both the intervention and control groups. Multivariate linear regression models were used to determine the effect of CPIPE on the outcome measures, controlling for relevant confounders. In the model building process, we first included facility-level random components, but the ICC values were negligible. We thus present the results of the multivariate linear regression models with robust standard errors. A difference-in-difference analysis was used to estimate the attribution effect of CPIPE on outcome measures. For a given outcome, the intervention effect is the outcome change from baseline to endline for the intervention group minus that of the control group [48]. We used an intent-to-treat design, thus did not account for movement of providers. A p-value of 0.05 was considered statistically significant. Data were analyzed using STATA 15 [77].

### Qualitative analysis

IDI transcripts were analyzed using a codebook thematic analysis approach [78–80]. A coding framework, developed deductively based on the interview guide, was iteratively refined with the addition of inductive codes following coding of initial transcripts. A team of four researchers (MG, BO, EN, JK) coded using the coding framework in Dedoose version 8.3.19 [81], with discrepancies addressed by discussion. Codes were queried and analytical memos drafted to identify themes with illustrative quotes.

### Mixed methods integration

We used a convergent mixed-methods methods design, with the interview guide developed to help illustrate, expand, and explain the quantitative results [49]. IDI respondents were drawn from the survey sample to connect the findings. We present the quantitative and qualitative findings separately in the results section and then draw on both to interpret the findings in the discussion [82].

## Results

### Participant characteristics at baseline

The analytic sample is the 80 providers who completed both baseline and endline surveys. There were few differences in the demographic characteristics of participants in the intervention and control groups at baseline; but all, except marital status, were not statistically significant (Table 2). Most participants were female (65% of control and 80% of intervention), between 30 and 40 years of age (50% of control and 40% of intervention) and married (85% of control and 72% of intervention). Most were nurses or midwives (62.5% of control and 77.5% of intervention), and about a third had been providers for less than 5 years. Less than 3% in both groups reported ever participating in a formal training on stress management; 3% and 5% in the control group reported ever having a training in unconscious bias and interpersonal interactions with patients, respectively, compared to 15% for both trainings in the intervention group.

**Table 2:** Baseline characteristic of the study participants, N=80.

| Characteristics | Category | Control (N=40) | Intervention (N=40) | P |
| --- | --- | --- | --- | --- |
|  |  | Frequency (%) | Frequency (%) |  |
| **Gender** | Male | 14 (35.0) | 8 (20.0) | 0.133 |
|  | Female | 26 (65.0) | 32 (80.0) |  |
| **Age** | 30 or below years | 9 (22.5) | 12 (30.0) | 0.632 |
|  | 31-40 | 20 (50.0) | 16 (40.0) |  |
|  | 41 or above | 11 (27.5) | 12 (30.0) |  |
| **Current marital status** | Single | 2 (5.0) | 11 (27.5) | 0.003 |
|  | Currently Married | 34 (85.0) | 29 (72.5) |  |
|  | Divorced/Widow/separated | 4 (10.0) | 0 (0.0) |  |
| **Number of children** | No children | 1 (2.5) | 5 (12.5) | 0.149 |
|  | 1 to 3 | 26 (65.0) | 27 (67.5) |  |
|  | 4 or more | 13 (32.5) | 8 (20.0) |  |
| **Position** | Doctor | 2 (5.0) | 1(2.5) | 0.287 |
|  | Clinical officer | 3(7.5) | 0(0.0) |  |
|  | Nurse/Midwife | 25(62.5) | 31(77.5) |  |
|  | Support staff | 10(25) | 8(20.0) |  |
| **Years of work as a health provider** | 0 to 5 years | 12 (30.0) | 15 (37.5) | 0.169 |
|  | 6 to 10 years | 19 (47.5) | 11 (27.5) |  |
|  | More than 10 years | 9 (22.5) | 14 (35.0) |  |
| **Years in current facility** | 2 years or below | 22 (55.0) | 20 (50.0) | 0.654 |
|  | 3 or more years | 18 (45.0) | 20 (50.0) |  |
| **Days of work per week** | 5 or fewer | 31 (77.5) | 29 (72.5) | 0.606 |
|  | More than 5 days | 9 (22.5) | 11 (27.5) |  |
| **Training on how to deal with stress** | No | 39 (97.5) | 39 (97.5) | 1.000 |
|  | Yes | 1 (2.5) | 1 (2.5) |  |
| **Ever had training on unconscious bias (n=79)** | No | 38 (97.4) | 34 (85.0) | 0.108 |
|  | Yes | 1 (2.6) | 6 (15.0) |  |
| **Training on interpersonal interactions** | No | 38 (95.0) | 34 (85.0) | 0.263 |
|  | Yes | 2 (5.0) | 6 (15.0) |  |
| **Highest level of education** | Secondary or below | 8 (20.0) | 10 (25.0) | 0.847 |
|  | College (middle level) | 26 (65.0) | 25 (62.5) |  |
|  | University or above | 6 (15.0) | 5 (12.5) |  |
| **Perceived social status of family growing up** | Bottom half | 25 (62.5) | 28 (70.0) | 0.478 |
|  | Upper half | 15 (37.5) | 12 (30.0) |  |
| **Perceived social status of self now** | Bottom half | 18 (45.0) | 11 (27.5) | 0.104 |
|  | Upper half | 22 (55.0) | 29 (72.5) |  |
| **Social mobility** | Upward mobility | 27 (67.5) | 33 (82.5) | 0.163 |
|  | No change | 7 (17.5) | 6 (15.0) |  |
|  | Downward mobility | 6 (15.0) | 1 (2.5) |  |
| **From the County** | No | 10(25.0) | 18(45.0) | 0.061 |
|  | Yes | 30(75.0) | 22(55.0) |  |
| **Religion** | Catholics | 12 (30.0) | 12 (30.0) | 0.597 |
|  | Methodist/Presby/Anglican | 2 (5.0) | 3 (7.5) |  |
|  | Protestant/Pentecostal | 6 (15.0) | 10 (25.0) |  |
|  | Seventh Day Adventist | 20 (50.0) | 15 (37.5) |  |

The full sample consists of 80 providers (40 each in the intervention and control groups). Total less than 80 due to missing data are noted.

### Quantitative results on impact of intervention on study outcomes

There were no statistically significant differences between the intervention and control groups in any of the key outcomes at baseline. Bivariate analysis showed slight increases from baseline to endline in stress and unconscious bias knowledge scores, as well as decreases in perceived stress and burnout scores in the control group, but these changes were not statistically significant (Table 3). There were, however, statistically significant increases in stress and implicit bias knowledge scores and decreases in stress and burnout levels in the intervention group (Table 3). The average stress knowledge score increased from 7.8 to 9.5, while implicit bias knowledge increased from 8.9 to 10.7 (p<0.001) for the intervention group. On the other hand, perceived stress scores decreased from 20.9 to 18.6 (P<0.01), while burnout scores decreased from 3.6 to 3.0 for the intervention group (p<0.001). There were decreases in the explicit bias scores for both groups, but the magnitude of change was descriptively, though not significantly, larger in the intervention group. The mean score on the low SES vignette decreased from 17.4 to 15.7 (p=0.05) in the intervention group compared to 16.30 to 14.9 (p=0.08) in the control group. Mean scores for the high SES vignette also decreased from 17.9 to 15.3 (p=0.006) in the intervention group compared to 17.0 to 15.0 (p=0.01) in the control group. IAT scores decreased for both groups, with a larger and statistically significant difference in the control group (p=017). Self-reported provision of PCMC increased from 57.8 to 65.5 (p=0.03) in the intervention with no statistically significant change in the control group. There were no significant changes in HRV from baseline to endline for both intervention and control groups. The average hair cortisol level however increased in the control group (p<0.05) but decreased in the intervention group (p>0.05).

**Table 3:**
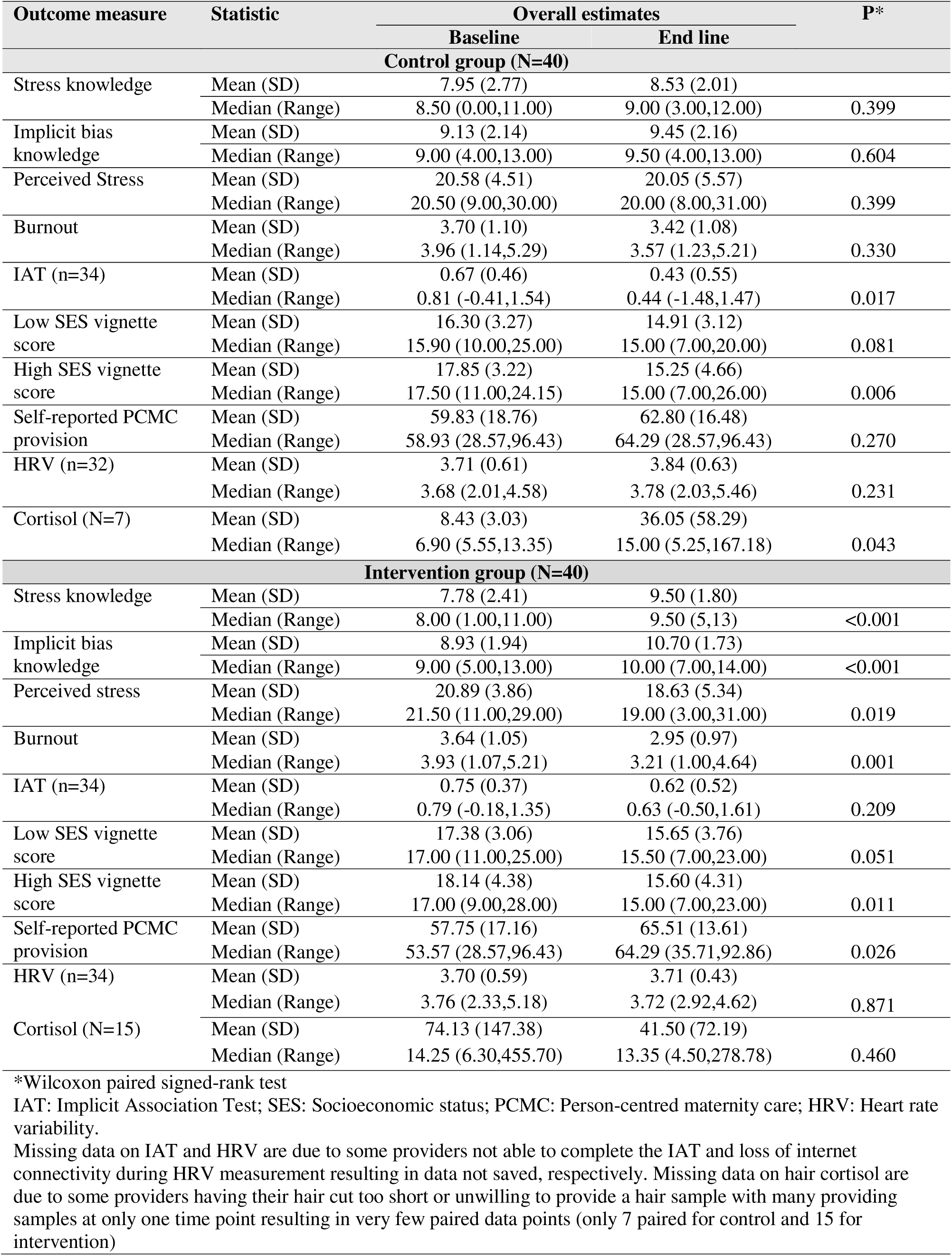
Estimates of primary outcomes at baseline and endline for control and intervention groups.

In multivariate analysis, controlling for relevant confounders (Table 4), these relationships persisted with greater and statistically significant increases in knowledge scores, decreases in stress, burnout, and explicit bias scores, and increases in self-reported PCMC from the baseline to endline in the intervention group, with smaller or no statistically significant changes in the control group. For example, controlling for other factors, perceived stress scores decreased by 2.5 points from baseline to endline in the intervention group (/3=-2.5; CI:-4.70, -0.30), with no statistically significant change in the control group (/3=-0.76; CI:-2.99,1.47). Similarly, self-reported PCMC provision increased by 8 points in the intervention group (/3=-8.07; CI:1.29,14.84), with no significant change in the control group (/3=-1.71; CI:-5.40,8.82). The change in IAT scores also remained marginally significant in the control group (/3=- 0.25; CI:-0.49, -0.01), but not significant in the intervention group (/3=-0.05; CI:-0.27,0.17). Differences in HRV and hair cortisol levels were not statistically significant. In difference-in-difference analysis (Table 5), only the percentage difference in implicit bias knowledge was significant, with 36% of the change attributable to the intervention. Although not statistically significant, the intervention accounted for about 81% of the decrease in perceived stress and about a third of the decrease in burnout and explicit bias and increase in PCMC provision.

**Table 4:** Multivariate analysis of primary outcomes stratified by study group.

| Study period | Control |  | Intervention |  |
| --- | --- | --- | --- | --- |
|  | Coefficient (95 % CI) | P | Coefficient (95 % CI) | P |
| **Stress knowledge** |  |  |  |  |
| Baseline | Reference | - | Reference |  |
| End line | 0.66 (-0.23,1.55) | 0.144 | 1.59 (0.75,2.43) | <0.001 |
| **Implicit bias knowledge** |  |  |  |  |
| Baseline | Reference | - | Reference |  |
| End line | 0.28 (-0.63,1.18) | 0.543 | 1.64 (0.82,2.45) | <0.001 |
| **Perceived stress** |  |  |  |  |
| Baseline | Reference | - | Reference |  |
| End line | -0.76 (-2.99,1.47) | 0.498 | -2.50 (-4.70, -0.30) | 0.026 |
| **Burnout** |  |  |  |  |
| Baseline | Reference | - | Reference |  |
| End line | -0.48 (-0.95,-0.02) | 0.041 | -0.82 (-1.24, -0.41) | <0.001 |
| **IAT** |  |  |  |  |
| Baseline | Reference | - | Reference |  |
| End line | -0.25 (-0.49,-0.01) | 0.042 | -0.05 (-0.27,0.17) | 0.652 |
| **Low SES vignette score** |  |  |  |  |
| Baseline | Reference | - | Reference |  |
| End line | -1.55 (-3.01,-0.08) | 0.039 | -2.09 (-3.59,-0.58) | 0.007 |
| **High SES vignette score** |  |  |  |  |
| Baseline | Reference | - | Reference |  |
| End line | -2.79 (-4.78,-0.79) | 0.007 | -3.14 (-5.15,-1.12) | 0.003 |
| **PCMC** |  |  |  |  |
| Baseline | Reference | - | Reference |  |
| End line | 1.71 (-5.40,8.82) | 0.633 | 8.07 (1.29,14.84) | 0.020 |
| **HRV** |  |  |  |  |
| Baseline | Reference | - | Reference |  |
| End line | 0.21 (-0.07,0.49) | 0.146 | 0.01 (-0.21,0.23) | 0.922 |
| **Cortisol** |  |  |  |  |
| Baseline | Reference | - | Reference |  |
| End line | 25.52 (-6.97,58.01) | 0.101 | -39.66 (-98.24,18.93) | 0.178 |

Notes:
1. Stress knowledge model controls for sex, social status, length of provider and being a clinician 2. Implicit biased knowledge model controls for education level and length of provider 3. Perceived stress model controls for marital status, role, and work hours 4. Burnout model controls for sex, age, religion, social status, and work hours 5. IAT model controls for marital status, days per week worked, and being a clinician 6. Low SES model controls for hours worked and being a clinician 7. High SES model controls for hours worked and being a clinician 8. PCMC model controls for education level, religion and being a clinician 9. HRV model controls for sex and length of provider 10. Cortisol model controls for length of provider, hours worked and being a clinician

**Table 5:**
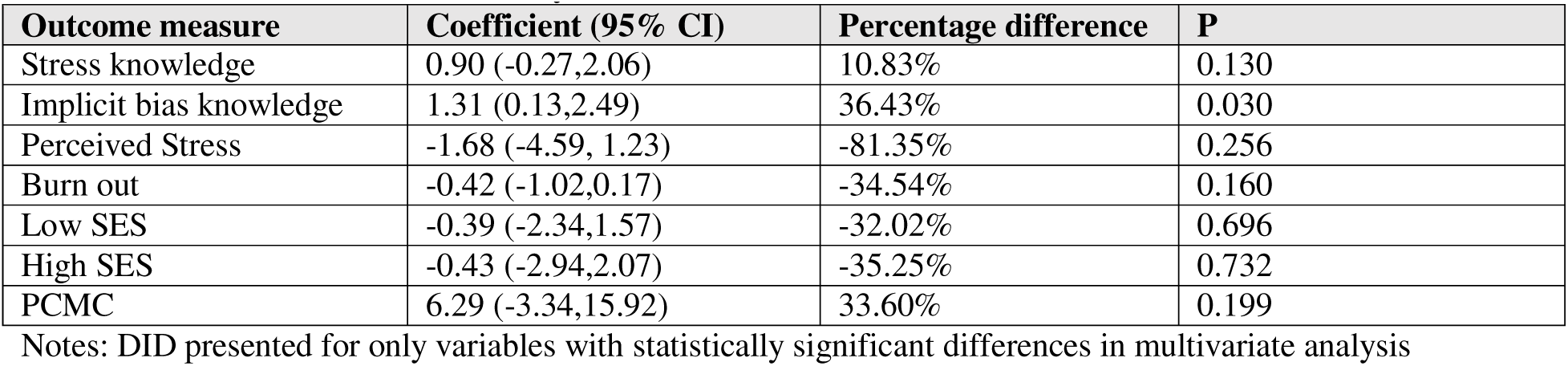
Difference in difference analysis of CPIPE effect on different outcomes.

### Qualitative results

We present themes based on the Social Cognitive Theory constructs—impact on knowledge, skills, self-efficacy, behavior, experience, and environment related to the intervention outcomes—organized by the Ecological Framework’s multiple levels of influence—individual, interpersonal, and institutional levels. Provider accounts of the intervention impacts, however, usually cut across different constructs and levels as reflected in the quotations below and in Table 6.

**Table 6:**
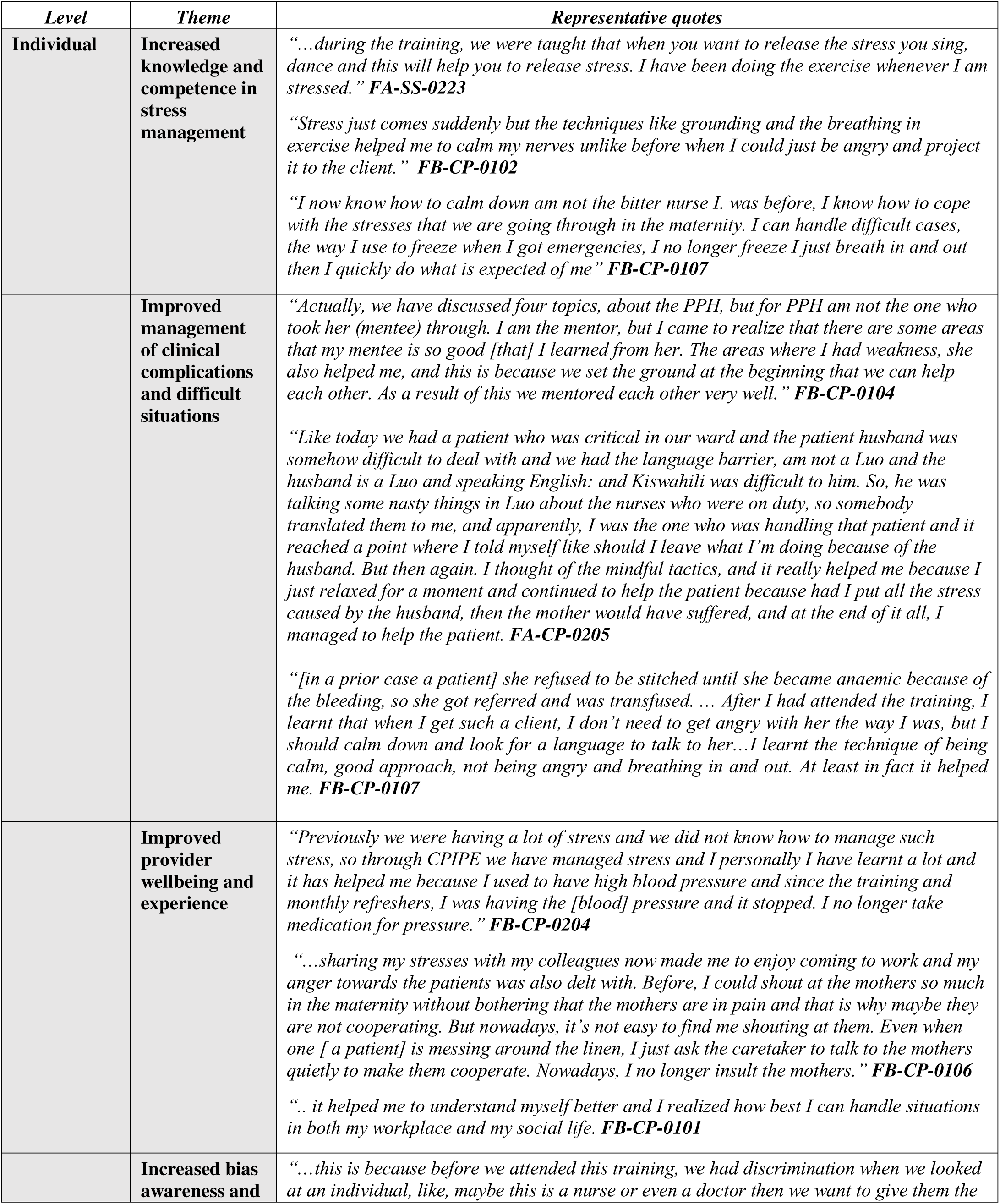
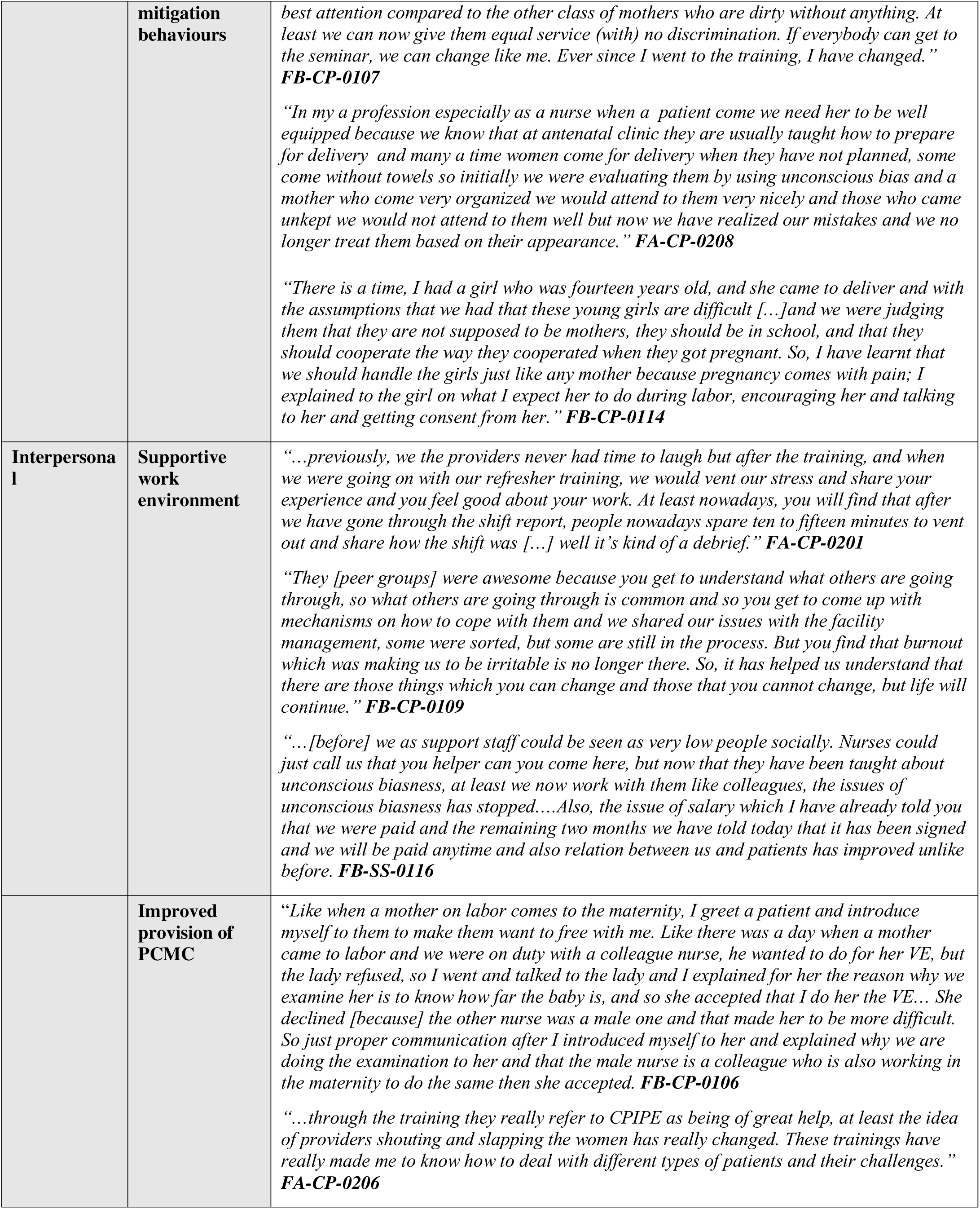
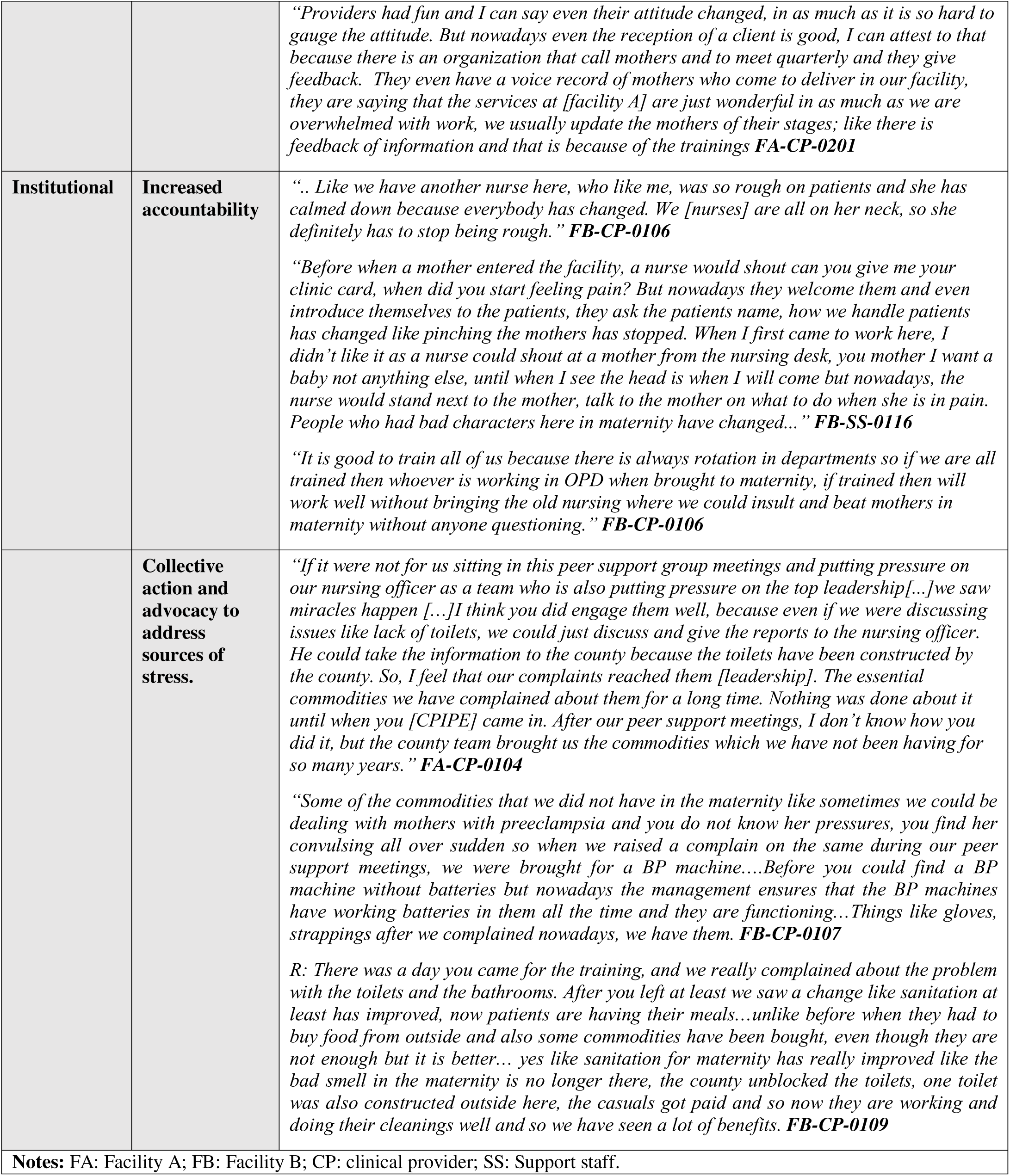
Intervention Impact themes and representative quotes.

### Individual level Impacts

At the individual level, participants credited the CPIPE intervention for increasing their knowledge and competency in stress management, bias mitigation, and PCMC provision, which in turn increased their confidence, attitudes, and behaviors, resulting in positive impacts on their personal wellbeing and experiences and patient experience. IDs for quotations represents the facility (FA and FB: facility A and B) and provider type (CP: Clinical Provider and SS: Support staff). These are used to avoid potentially identifying providers.

### Increased knowledge and competence in stress management

The training content on stress management was particularly salient and credited for increasing providers’ ability to cope with and manage stress. Participants discussed how increased knowledge of different stress management techniques contributed to reductions in their stress level.

> *“…whatever I was taught during training has been of great help to me, because initially I had a lot of stress, and when I attended the training, I was taught how to manage the stress that I may encounter; as we talk now, the stress level has reduced and now I am good.” **FA-SS-0222***

Increased knowledge of stress management was reflected in participants’ descriptions of the different techniques they were using to manage stress. These included talking to a trusted person, singing, dancing, self-care activities, breathing, and other mindfulness-based techniques.

> *“…the topic of stress really helped me because sometimes I could get so stressed, but I could not tell anyone. So, I came to realize that [in] sharing with a colleague or a person whom I trust, then I can get relieved.” **FB-CP-0106***

### Improved management of clinical complications and difficult situations

Further, training and mentorship were credited with increasing competence in the clinical management of obstetric emergencies such as post-partum hemorrhage (PPH) and birth asphyxia, both previously reported major stressors. This in turn helped improve providers’ confidence when managing emergencies, leading to decreased fear and anxiety, which improved both clinical and person-centered care during emergencies.

> *“It helped me a lot because whenever I have any stress in the* wards*, then I know how to cope with, especially PPH. The anxiety that I had when I got a PPH at least I can now handle it so well and attend to it properly without fear.” **FB-CP-0107***

Providers also reported how the training enabled them to better manage difficult situations, which previously was cited as a reason for disrespect and abuse of patients, contributing to improvements in their interactions with patients in such situations. Increased confidence to manage complications and difficult situations were credited for calming both the provider and the patient.

> *“I have really* learnt *a lot; before attending the trainings, I did not know how to handle difficult patients and mothers in labor. And when I attended the training, I was trained how to handle such difficult cases.” **FA-SS-0223***

### Improved provider wellbeing and experience

Participants expressed a positive change in their wellbeing due to a reduction in stress, which they credited to the intervention activities. Some even noted changes in their health following the intervention.

> *“I benefited because I was so stressed, I could sit down and just get rumbles in my stomach, but when I attended the training, I got to know how to control my stresses and it really helped me.” **FB-SS-0115***

Others recounted how stress reduction techniques helped with managing their anger, which previously would have been directed towards patients. Providers reported higher job satisfaction because of the intervention which made them enjoy coming to work and improved their interactions with patients.

> *“… people* like *me we opened up and we got help from colleagues, something which I did not know that was so easy, that I could be feeling much lighter and happier coming to work.…Sharing my stresses with my colleagues now made me to enjoy coming to work and my anger towards the patients was also delt with.” **FB-CP-0106***

### Increased bias awareness and mitigation behaviors

The intervention was also credited for increasing providers’ awareness of implicit and explicit bias. This encouraged providers to reflect on their assumptions, which enabled them to control the impact of their biases and avoid discriminatory practices. For example, providers noted improved behaviors including avoiding discrimination based on their perceptions of a patient’s social status or appearance, a common practice before the intervention.

> *“So, initially we* were *evaluating [patients] by using unconscious bias and a mother who come very organized we would attend to them very nicely; and those who came unkept, we would not attend to them well. But now we have realized our mistakes and we no longer treat them based on their appearance.” **FA-CP-0208***

Others reported how the training on bias had enabled them to be more open-minded and empathize with their patients, without discriminating based on patient attributes such as ethnicity and age.

> *“Mostly* understanding *my clients because when I came to work in this facility, I was so negative. I just already had an opinion with the Kurias (an ethnic group) that they do behave like this or that. So, now am getting to understand them, and it has also helped me to open an avenue where I can begin a conversation; I can understand my clients and my colleagues other than sticking to my opinions that these people are like this, primitive and violent, which may not be so.” **FB-CP-0109***

## Interpersonal level Impact

At the interpersonal level, the intervention was credited for improving relationships among providers and between providers and patients leading to a supportive work environment and improved PCMC.

### Supportive work environment

Participants felt that the intervention made them more collaborative and open with colleagues. They also felt the intervention led to more friendly interactions with their colleagues. These facilitated a more supportive work environment, which further improved experiences at work.

> “*It made me* more *open to my colleagues, I listen more and when somebody is talking, I let them talk even if I was rushing somewhere, unless it was an emergency. I give them a listening ear. I spare more time for my colleagues.” **FA-CP-0201***

In addition, the mentorship intervention facilitated a non-hierarchical cross-cadre learning which enabled providers to enhance their clinical skills in a supportive environment. The peer support groups also provided an opportunity to debrief, share experiences at work and beyond, and discuss solutions together. These activities also enabled providers to better understand each other.

> *“We could discuss the challenges that we experience both at work and out of work then we could brainstorm on the solution to these challenges, we could do some activities like singing and dancing and this I saw to have helped us in change of mind set, you become relieved.” **FB-CP - 0110***

Improved relationships occurred within and across cadres. Support staff noted how clinical providers now treated them with more respect. Participants also reported improved relationships with their supervisors, who also noted improvements in their staff. For instance, one maternity-in charge discussed how due to the training she now listens more to the staff, which has improved her understanding of issues they are dealing with and improved their interactions.

> *“I have had staffs who have had issues sometimes, but due to such a training, I have been able to give them time to explain where they have been having issues and looking on sharing basis on how best to handle certain situations. Like in a* situation *where someone failed to report on duty, I ask them to explain what happened and after explanation now I am able to understand that someone can be in such a situation and I am able to tell them that they are able to communicate when caught in such situations and so she appreciated because we did not take any disciplinary measures against her. We understood the situation and… I placed someone to work for her and the issue ended well. Unlike before when I could forward the case for a disciplinary action before listening to the staff. **FB-CP-0101***

### Improved provision of PCMC

The intervention was credited for influencing providers’ approach to patient care. Providers noted how the intervention had impacted general patient-provider interactions by improving providers’ knowledge in communicating with patients, friendly service provision, and compassionate care.

> *“After the trainings that I* received*, I was able to handle and talk to women in a friendly way without being biased, and with that the women were getting quality care from me. And during delivery, I could talk to women in a soothing way until the baby is delivered and they all go alive.” **FA-CP-0222***

As seen in previous quotes, improved individual provider experience was reported to translate into better interactions with patients thus improving PCMC. Providers shared that these positive changes were appreciated by their patients, leading to more trust, positive perceptions from the community, and increased service utilization.

> *“.. it has come to a point* where *community people have started saying that [Facility B] is good, and they no longer go to private facilities. They are saying that ‘here we are treated well, nobody is becoming rough’.” **FB-CP-0106***

## Institutional level Impacts

At the institutional levels, the intervention was credited for improving accountability and advocacy with leadership which served as reinforcement and helped to address some sources of stress. Impacts on all levels led to a more positive facility culture which created an enabling environment for ongoing behavior change.

### Increased accountability

Because most providers in the maternity unit participated in the training, they were holding each other accountable by calling each other out when they behaved out of line. Further, changes in most providers also motivated even potentially resistant providers to change their behaviors. Providers noted reminding each other of their biases to avoid discriminatory care in their facilities.

> *“[The] training on* unconscious *bias, the way somebody appear, you could attend to somebody based on how they look but with time, now that everybody was trained, now when unconscious (comes-up), then your colleagues are able to remind you of the unconscious [bias] training and then you get back to your conscious [self]; you think that I should treat everybody the same. The training has really helped.” **FA-CP-0201***

### Collective action and advocacy to address sources of stress

Many felt the intervention strategy to engage leadership made it easier to implement the project activities and enhanced support for the intervention. Leadership engagement, together with the peer support group strategies, were credited for facilitating collective action and advocacy to address health systems issues including lack of supplies, poor infrastructure, and delayed salaries.

> *“You saw it here in our meeting, we had so many challenges but so many of the challenges had been sorted. Like we had a challenge of salaries for casuals, had it been that we did not engage the county then we could have not paid them… it’s because of the study. The issues of supplies you had it mentioned in the meeting. The issue of* pharmaceutical *and non-pharm came out loudly in our last meeting and out of that meeting, it never took like one month before we could see the supplies trickling into our facility, so I think that leadership engagement has really brought out things to….be done in the right way.” **FA-CP-0101***

## Discussion

The findings from our mixed methods evaluation shows that the CPIPE intervention contributed to increases in provider knowledge and competency in stress management and bias mitigation, decreases in stress, burnout, and explicit bias levels, and increase in self-reported PCMC. These findings are consistent with our hypothesis. Although there were small changes in the control group, likely due to socially desirable responding, repeated testing, and maturation [48], the greater changes in the intervention group provide evidence for improvements due to the intervention. Further, the difference-in-difference analysis suggests that about a third or more of the changes may be due to the intervention, but the small sample size limited our ability to achieve significance levels for attribution effects. The decrease in average hair cortisol levels in the intervention group compared to an increase in the control group (although not statistically significant likely due to the small sample size), also suggests a potential physiologic impact of the intervention on provider stress levels. Finally, although there were no significant changes in IAT scores in the intervention group, the qualitative data highlights providers’ awareness of their biases and adoption of strategies to mitigate the effects of those biases. Qualitative data also illustrate mechanisms by which the intervention impacted the other study outcomes.

The reduction in provider stress and burnout levels is a critical finding, given the intervention did not directly address stressors. One potential pathway for this effect is that the intervention influenced how providers perceived these stressors. It is recognized that people exposed to the same stressors may have different physiological and psychological responses, depending on how they perceive the stressors [22]. Further, the training provided participants with knowledge and tangible skills to cope with stress and prevent burnout. Thus, although stress is unavoidable in the life of healthcare workers due to the nature of their work, the impact of stressors can be diminished by equipping them with the tools necessary to change their perception and manage the stressors and promoting positive coping mechanisms. Qualitative data also show that the intervention contributed indirectly to addressing some of the stressors, lessening the frequency of exposure to stressors. Prior interventions to prevent burnout among providers have leveraged similar mechanisms including individual-focused strategies (such as stress management training, self-care workshops, mindfulness, etc.) as well as structural approaches (e.g., workload or schedule changes and improving workplace environment) [83].

The decreased explicit bias scores are likely due to the increased knowledge of bias, which qualitative data show was being translated into action to avoid discrimination and provide PCMC to all patients regardless of their social status. The non-significant decrease in IAT scores in the intervention group, on the other hand, reflects the implicit bias training goal of increasing awareness and mitigation behaviors, rather than removing the underlying associative process [84]. Additionally, short term implicit bias trainings are insufficient to root out deeply held biases. Instead, the goal is to raise people’s awareness of implicit bias to facilitate the development of structural changes to mitigate the effects of individual biases [41,85]. The significant reduction in IAT scores in the control group was, however, unexpected; although likely due to the low test-retest reliability of the IAT [86,87]. Prior studies have shown lower subsequent IAT scores regardless of intervention exposure, calling for caution with repeated IAT measures [86,87]. The CPIPE intervention included several components recommended for integrating implicit bias recognition and management into health professions curricula, including, creating a safe and nonthreatening learning context, increasing knowledge about the science of implicit bias, emphasizing how implicit bias influences behaviors and patient outcomes, increasing self-awareness of existing biases, improving conscious efforts to overcome implicit bias, and enhancing awareness of how implicit bias influences others [88]. These likely contributed to the positive effects of the intervention with evidence that providers were more aware of their biases and adopting strategies to mitigate their effects. Increased bias awareness is a first step towards mitigation. Further, providers were learning how to replace biased responses with responses more consistent with their goals, which can help mitigate the effects of bias [85]. Emerging research also suggest that it is possible to reduce the effects of people’s bias through activities that elevate the alternate selves and goals that people endorse, without actually removing their deep-seated associative biases—referred to as sidelining implicit bias—due to the inherently complex and often contradictory nature of people [89,90].

The CPIPE outcomes are a result of the synergistic effects of the different intervention strategies. As posited in the social cognitive theory, people can change their behavior despite obstacles if they feel capable and confident in their ability to engage in the behavior (self-efficacy), believe the change benefits them (outcome expectancies), and if there are tangible goals, positive role models, and reinforcement [45]. Further, learning and behavior change occurs in a social context with a dynamic and reciprocal interaction of the person, environment, and behavior [42]. Training contributes to self-efficacy and outcome expectancies, and is necessary, but often insufficient by itself to achieve and sustain behavior and facility culture change. Yet, most interventions on PCMC continue to focus on only training, with most content focused on only respectful maternity care [91]. CPIPE is unique because, in addition to the unique training content and delivery, the intervention package included theory and evidence-based multilevel integrated strategies, which played a critical role in the success of the intervention.

Peer support programs, while quite common among patient groups, are not as widely utilized among healthcare workers in SSA [92–97]. The cadre-specific in-person peer support groups provided an opportunity for maternity providers to offer emotional and social support, share knowledge and experience, practice coping mechanisms to reduce stress, and discuss ways to address bias and provide PCMC. Mentoring on the other hand an accepted approach to support ongoing professional development guided by mentees learning needs [98–100]. The CPIPE onsite mentorship program created new relationships between providers contributing to positive environment for providing compassionate care, while improving providers’ competencies to manage clinical emergencies. Embedded champions are recognized as central to the success of implementation and behavior change [101–103]. CPIPE embedded champions served as trainers, mentors, conveners, liaisons, role models, and advocates, which contributed to the intervention success, increased self-efficacy of other providers, and provided reinforcement for behavior change [42]. Finally, leadership engagement is critical for successful implementation, ownership, and sustainability of any intervention [104]. In CPIPE, leadership engagement from the onset contributed to county buy in, facilitated implementation, and granted to access leadership such that concerns raised at peer support meetings could be brought up to management for resolution. The reciprocal interactions of individual behavior, peers, mentors, mentees, embedded champions, and leaders thus all created an enabling environment for learning and reinforcing behavior change.

Our findings have several implications. First, given increasing documentation of stress and burnout among healthcare workers [27,29,31,105], CPIPE provides an example of a theory and evidence-based approach to prevent burnout, promote the wellbeing of providers, and improve quality of care. Second, given the role of bias in healthcare inequities and the dearth of evidence on effective ways to address it [15,17,33], the CPIPE intervention will be valuable in efforts to promote health equity through bias awareness and mitigation. Third, there is still a major gap in theory and evidence-based interventions to promote responsive, compassionate, and respectful maternity care [91,106]. This work thus extends the limited research on interventions to improve PCMC, with the added strength of addressing equity and provider wellbeing and experience. Finally, CPIPE provides an example of how to facilitate a supportive workplace culture that both motivates and holds providers accountable for specific behaviors. Planning for and creating policies within health systems for the implementation of such interventions could thus have multiplicative effects beyond the outcomes of this study.

## Limitations and strengths

First, given this was a pilot study with a small sample in one setting to assess feasibility, generalizability is limited. The small sample size also likely contributed to the non-significant difference-in-difference results. Second, individual provider randomization was not appropriate given the facility level activities and high potential for contamination. Thus, we utilized a cluster design; but exact matching of facilities was not possible due the highly variable size of the facilities in the study county. This resulted in some differences between the intervention and control sites. Third, self-reported measures are prone to social desirability bias. We included the IAT and physiologic measures of stress to address this limitation, but the low test-retest reliability of the IAT and acute nature of HRV (influenced by what is happening in the moment) limited their utility, while people’s reluctance to have their hair cut significantly reduced the analytic sample for hair cortisol. Finally, although the goal is to improve patient experience, patient interviews were not included in this pilot phase and the one-time assessment period at the end of the intervention limits our ability to assess sustained effects.

Nonetheless, the study has several strengths. To our knowledge, this is the first study to show the effect of an integrated provider targeted intervention on perceived stress, burnout, and bias—key drivers of inequities in PCMC. The findings address a gap in efforts to improve both patient and provider experience as well as advance health equity. Second, despite limitations of not being able to randomize individually, the pretest-posttest control group design allowed us to address common threats to internal validity, thus increasing the internal validity of the results [48]. We also used validated measures to assess our outcomes further increasing internal validity. Despite the limitation of not including patient interviews, the self-reported PCMC results are promising as prior research has shown that provider reports in this setting are close enough to patient reports, despite the danger of social desirability bias [13,14,16]. Finally, the convergent mixed-methods methods approach provided a more nuanced understanding of the impact of the intervention. The success of this pilot sets it up for testing in a larger sample with patient reported experience measures and outcomes and with longer term follow-up to strengthen the evidence for effectiveness, generalizability, and sustainability.

## Conclusion

CPIPE is a theory and evidence-based integrated provider intervention that addresses key interrelated drivers of poor PCMC—provider stress, burnout, and implicit and explicit bias—as a way of improving and achieving equity in PCMC. Through this pilot, we have demonstrated the preliminary effectiveness of CPIPE. The evaluation results show strong promise of its ability to reduce provider stress and burnout, as well as to mitigate the effects of explicit and implicit bias to improve PCMC for the most vulnerable.

Further, the multilevel intervention strategies contributed to individual provider and facility level changes that created an environment conducive for providing responsive, respectful, and compassionate maternity care. The findings from this study have great promise for other settings seeking to improve both provider and patient experience, while centering the experience of the most vulnerable patients. Future studies in more diverse settings will strengthen the evidence to inform programs and policies on such interventions.

## Data Availability

All data produced in the present study are available upon reasonable request to the authors

## Abbreviations

CPIPE: Caring for providers to improve patient experience
PCMC: Person-centered maternity care
SES: Socioeconomic status
IAT: Implicit association test
HRV: Heart rate variability
FA: Facility A
FB: Facility B
CP: clinical provider
SS: Support staff.

## Author contributions

PAA conceptualized and designed the study. BAO, ENO collected the data. JO, MG, BAO, ENO, JK, PAA contributed to data analysis and interpretation. All authors contributed to study implementation and writing of the manuscript. All authors approved the final version of the manuscript for submission.

## Acknowledgements

We thank the Migori County, sub-county, and health facility leadership, and the providers who participated in the study. We are especially grateful to the four CPIPE Embedded Champions (Joash Mayuya, Mary Khaemba, Gladys Nyaberi, and Beatrice Bange), who facilitated activities at their facilities.

